# Association between potentially inappropriate medication use and all-cause mortality among older adults with dyslipidemia: a nationwide cohort study

**DOI:** 10.1101/2025.11.23.25340838

**Authors:** Sungju Lee, Seungbong Han

## Abstract

**Background:** Potentially inappropriate medications (PIMs) are known to be associated with adverse outcomes in older adults, yet evidence among those with dyslipidemia—who often experience polypharmacy—remains limited.

**Methods:** We conducted a nationwide, retrospective cohort study in South Korea using the Health Insurance Review and Assessment Service (HIRA) claims linked to national death records. Adults aged ≥65 years with ≥2 dyslipidemia diagnoses between January and June 2018 were classified as exposed to PIMs per the Drug Utilization Review (DUR) list and followed from a landmark date (July 1, 2018) through June 30, 2024 for all-cause mortality. The primary objective was to compare mortality between the PIM and non-PIM groups; comparisons were conducted using an unadjusted Cox model, an adjusted Cox model, and a propensity score–matched (PSM) cohort (propensity scores estimated via logistic regression including all baseline covariates). Balance was assessed using standardized mean differences, and proportional hazards were checked with scaled Schoenfeld residuals.

**Results:** Of 943,332 participants, 1.7% received at least one potentially inappropriate medication (PIM) at baseline. Over 6 years, the Kaplan–Meier cumulative mortality was 14.2% in the PIM group versus 11.9% in the non-PIM group (absolute risk difference, 2.3 percentage points; relative risk, 1.19). PIM exposure was significantly associated with increased six-year all-cause mortality risk in both propensity score–matched (HR, 1.12; 95% CI, 1.05–1.19) and multivariable-adjusted analyses (HR, 1.10; 95% CI, 1.05–1.15; both p < 0.001). In subgroup analyses, compared with a single PIM, multiple PIMs showed a numerically higher but non-significant risk (HR, 1.11; 95% CI, 0.79–1.56).

**Conclusions:** Among older adults with dyslipidemia, PIM prescriptions were associated with a 12% increased risk of all-cause mortality in the PSM cohort. These findings support cautious prescribing and regular medication reviews to minimize risk in this high-risk population.

## Introduction

### Background and Rationale

South Korea has the fastest-aging population among OECD member nations, with the share of people aged 65 and over surpassing 20% in 2024, accounting for one-fifth of the total population, and it is projected to exceed 40% by 2050[1, 2]. With this demographic shift, the utilization rate of medical services is expected to rise[3], which, in turn, is anticipated to raise both the extent of medication use among older adults and the risk of inappropriate prescriptions[4, 5].

Adults aged 65 and over have a higher burden of chronic diseases and disabilities, making them more likely to take medications compared to younger age groups, and cases of taking multiple drugs simultaneously to treat underlying conditions are common[4]. However, with advancing age in older adults, the reserve capacity of cells, organs, and body systems declines, renal function decreases, and changes in fat tissue distribution occur, which can affect the accumulation and metabolic pathways of certain drugs. Moreover, a combination of factors—including reduced homeostasis, frailty, and inadequate nutritional status—renders them more vulnerable to drug interactions[6, 7]. These factors ultimately affect the health and survival of older adults.

To address these issues, the American Geriatrics Society (AGS) first published the AGS Beers Criteria^®^ in 1991 and, through its sixth revision in 2023, has specifically identified potentially inappropriate medications (PIMs) for adults aged 65 and older, classifying them as priority drugs to be avoided[8]. Furthermore, many countries, including those in Europe, have established other explicit criteria such as the STOPP/START Criteria to periodically review lists of medications requiring caution in older adults, and have developed strategies to reduce inappropriate prescriptions by reflecting each country’s characteristics and the availability of medications[5].

The Ministry of Food and Drug Safety (MFDS) of South Korea, taking into account domestic circumstances and referencing international criteria such as the Beers Criteria, McLeod’s list, and STOPP/START Criteria, published the “A Guide to Proper Medication Use in Older Adults” in 2009 and 2015, respectively[9]. Since 2015, the Health Insurance Review and Assessment Service (HIRA) has been using the Drug Utilization Review (DUR) system to monitor medications prescribed to patients aged 65 and over, reviewing and blocking inappropriate prescriptions before dispensing[10]. In addition, since 2023, a new indicator called the “Prescription Rate of Medications Requiring Caution in Older Adults” has been introduced in the drug benefit quality assessment to help prevent the prescription of PIMs[11].

However, while current domestic and international guidelines provide detailed recommendations across various conditions—including dementia, cognitive impairment, heart failure, and urinary incontinence—clear prescribing guidance specifically for older adults with dyslipidemia remains lacking.

Dyslipidemia is highly prevalent among older adults, with U.S. data showing 34.7% having total cholesterol ≥200 mg/dL and Korean data indicating prevalence rates exceeding 50% in those in their 60s[12, 13]. It not only contributes to atherosclerosis but is also associated with greater susceptibility to ischemic heart disease and stroke[14, 15]. Moreover, most treatments for dyslipidemia, including statins, require long-term administration and may cause adverse effects such as myalgia, liver injury, and impaired renal function[16, 17]. Among older adults with dyslipidemia, the presence of multiple comorbidities often increases both the number and types of prescribed medications, thereby heightening the chance of being prescribed PIMs[18]. Therefore, older adults with dyslipidemia represent a particularly vulnerable subgroup in terms of medication safety. Despite this, most prior research on PIMs has examined general older populations, with limited evidence focusing specifically on older adults with dyslipidemia.

Despite numerous studies on PIMs, no study has directly evaluated the impact of PIM exposure in older adults with dyslipidemia. A recent meta-analysis reported that increases in PIM use were linked to higher mortality among older adults[19], and similar trends have been observed in studies focusing on older adults with specific conditions such as dementia or kidney disease[20, 21]. In this context, there is a need for extended research on the mortality of patients with dyslipidemia resulting from PIM prescriptions. Continuous supplementation of explicit PIM criteria requires the accumulation of objective data and the establishment of solid evidence[5]. Given the relatively short history of PIM criteria, research investigating the long-term effects of PIM prescriptions in older adults with dyslipidemia could serve as an important foundation for safer and more effective medication prescribing in the future.

### Objectives

This study aims to compare six-year all-cause mortality between older adults (≥65 years) with dyslipidemia who were prescribed PIMs and those who were not, using multiple analytical approaches including multivariable Cox regression and propensity score–matched cohort analysis.

Specifically, this study seeks to analyze the specific components of PIMs prescribed to older adults with dyslipidemia, evaluate the relationship between the number of PIMs prescribed and mortality risk, and determine the impact of PIM prescriptions on long-term mortality outcomes using various statistical models. The findings will provide evidence supporting the importance of appropriate medication prescribing in the older adult population, raise awareness of adverse drug reactions, and serve as a foundational resource for updating the South Korean PIM list and improving prescribing guidelines. Furthermore, the results will inform indicator refinement in the HIRA’s drug benefit quality assessment.

## Materials and Methods

### Study Design

In this retrospective cohort conducted in South Korea, we estimated the impact of PIM prescribing on six-year all-cause mortality in older adults (≥65 years) with dyslipidemia. Baseline exposure assessment was conducted between January 1, 2018, and June 30, 2018, and participants were followed for mortality outcomes over six years until June 30, 2024. The analysis employed claims data extracted from the HIRA database, with mortality information obtained from death records of the Ministry of the Interior and Safety and the National Health Insurance Service.

### Data Source

The HIRA database includes claims from all healthcare providers reimbursed under the National Health Insurance program and Medical Aid programs, covering inpatient and outpatient services. The database captures nationwide claims from the provider types included in this study—tertiary hospitals, general hospitals, hospitals, long-term care hospitals, and clinics—for inpatient and outpatient services reimbursed under these two programs. Index treatment initiation dates were identified between January 1 and June 30, 2018. Diagnosis was coded using the Korean Standard Classification of Diseases, 7th revision (KCD-7; ICD-10–based); dyslipidemia was defined by E78.x codes. PIMs were defined using the HIRA “Medications Requiring Caution in Older Adults” list available through HIRA’s Drug Utilization Review (DUR) system. Mortality status was ascertained by linking death records from the Ministry of the Interior and Safety and the National Health Insurance Service to the HIRA database. The analytic dataset contained complete data on all variables included in the primary analyses. As a data-quality step, a targeted manual review verified the ages of nine participants aged ≥105 years; the recorded ages were confirmed to be accurate.

### Selection of Study Participants

Fig 1 shows the selection process for the analytic cohort. The study population was defined as older adults (≥65 years) who had at least two medical visits between January 2018 and June 2018 with dyslipidemia recorded as either a primary or secondary diagnosis code. This definition was based on previous studies that identified patients with dyslipidemia by setting a minimum of two visits as the inclusion criterion[22, 23].

**Fig 1.**
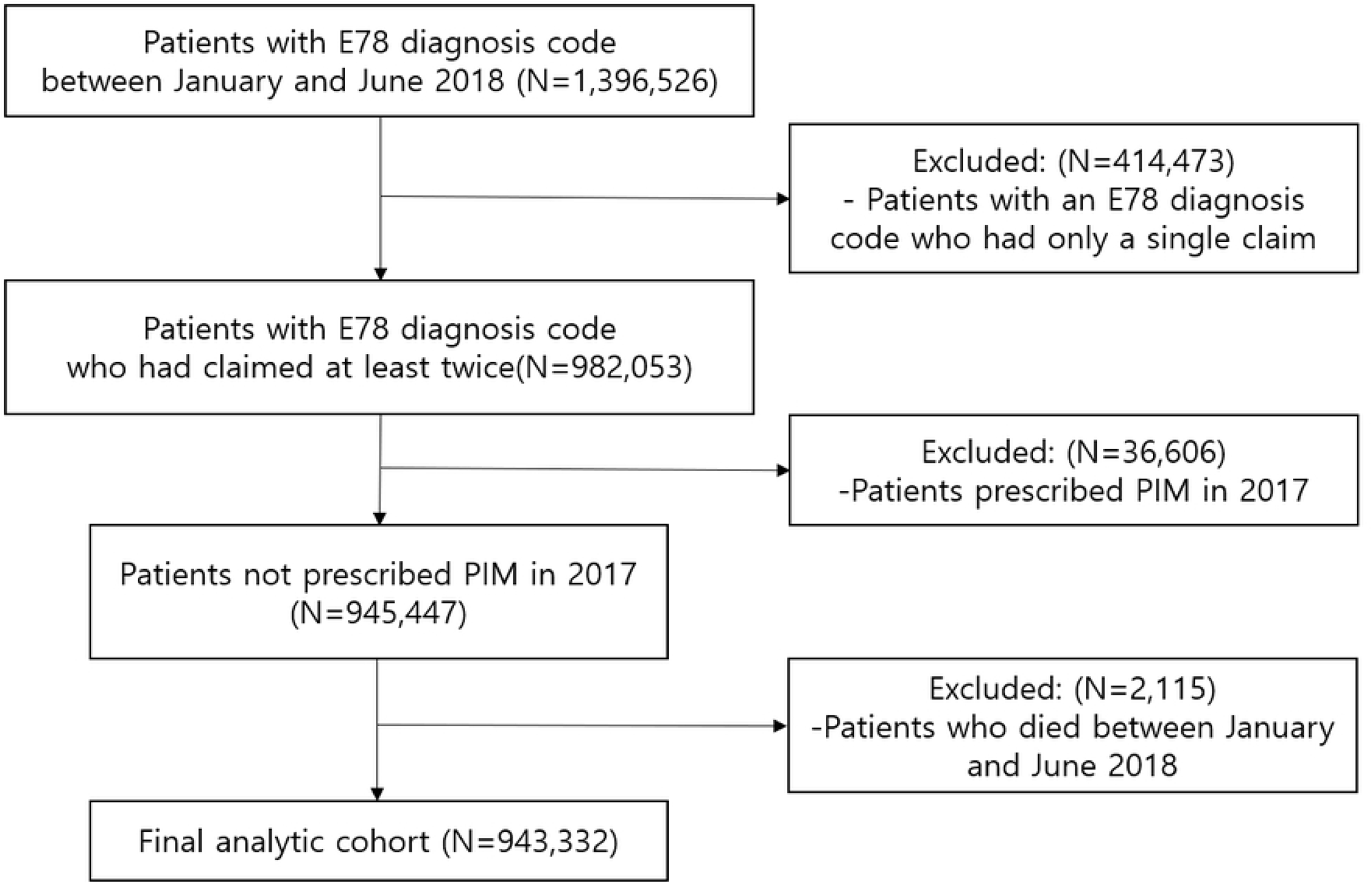
Flowchart of the Analytic Cohort Selection Process

Patients who had been prescribed PIMs during the year preceding the study start (January to December 2017) were also excluded to minimize potential confounding effects from prior medication exposure. Patients who died between January 1 and June 30, 2018, were excluded to avoid immortal time bias in the landmark analysis. The final analytic cohort comprised 943,332 patients meeting all inclusion criteria.

## Definition of Variables

### Main Exposure Variable

The primary exposure was whether a PIM was prescribed. In this study, PIMs refer to medications designated by the Ministry of Food and Drug Safety, with reference to international PIM lists, and monitored by the HIRA through its Drug Utilization Review (DUR) system. The DUR PIM monitoring list is announced monthly; in January and February 2018, 40 PIMs were designated, whereas from March to June, three drugs were removed from the list, leaving 37 PIMs as the target medications (The PIM list is provided in S1 Table). In early 2018, the national DUR PIM list was relatively narrow—focused mainly on benzodiazepines and tricyclic antidepressants—and may not reflect the broader PIM spectrum introduced in subsequent years.

### Outcome Variable

The outcome variable was all-cause mortality within six years. Mortality was tracked from the landmark date of July 1, 2018, to the end of the study period on June 30, 2024. We computed time-to-event, in years, as the interval from the landmark date to death for analysis.

### Covariates

Covariates included sex, age, type of insurance, type of medical institution, medical specialty, emergency treatment status, and inpatient/outpatient classification. Comorbidity variables were represented as binary variables for each of the 17 disease categories used in calculating the Charlson Comorbidity Index (CCI)[24]. A total of 17 disease categories were included, constructed using diagnosis records from claims statements within the six months preceding the index time (January 1, 2018). For the medical specialty variable, six specialties with a proportion of at least 1%—internal medicine, neurology, surgery, orthopedic surgery, neurosurgery, and family medicine—were treated as independent categories, while all other specialties were grouped into the “others” category for analysis (see S2 Table and S3 Table for detailed operational definitions).

## Statistical Methods

Continuous baseline variables were expressed as means (standard deviations), whereas categorical variables were presented as counts (percentages). Between the PIM-prescribed and non-PIM groups, categorical variables underwent analysis via Pearson’s χ² test, whereas continuous variables were tested using t-tests. Survival time accrued from the day after the landmark date. Participants without a recorded death date were administratively censored on June 30, 2024 (end of follow-up). We used the log-rank test to compare survival between groups; Kaplan–Meier estimates were generated for survival functions, and cumulative mortality incidence was graphically displayed. All variables in the primary analysis were complete with no missing data, eliminating the need for imputation procedures.

We first conducted unadjusted Cox proportional hazards regression to assess each variable’s association with mortality risk and to estimate the effect of PIM prescribing on all-cause mortality. Subsequently, three analytical models were constructed.

First, model 1 was prespecified based on a targeted review of five recent studies examining the association between PIM use and health outcomes[25–29]. Covariates were selected based on domain knowledge and a review of recent literature on PIMs and health outcomes—e.g., demographics, socioeconomic factors (such as race, education, and income level), baseline comorbidities (e.g., diabetes, hypertension, cardiovascular disease, rheumatologic disease, renal impairment), physical function (e.g., BMI, gait speed, frailty status), and medication burden—but inclusion was restricted to covariates that could be ascertained with sufficient validity from our claims data. Accordingly, model 1 included 11 covariates with reliable ascertainment in our claims data: age; sex; insurance type (a proxy for income level); myocardial infarction; congestive heart failure; peripheral vascular disease; cerebrovascular disease; rheumatologic disease; diabetes without complications; diabetes with complications; and renal disease. We did not adjust for baseline medication count because a valid concurrent medication measure at the index date could not be derived with sufficient accuracy. To reduce misclassification and avoid potential overadjustment, we prespecified its exclusion; baseline treatment intensity was partially captured by comorbidity burden.

Model 2 was developed using backward elimination, forcing inclusion of the PIM exposure variable. Variables were sequentially removed to minimize the Akaike information criterion (AIC), and the final set of retained variables is reported in the Results section. This approach allowed us to evaluate whether variables not emphasized in prior studies could provide additional explanatory power, and to retain only those covariates with the strongest statistical association, thereby improving model parsimony and statistical power.

Propensity score matching (PSM) was then implemented to minimize residual selection bias by establishing covariate balance between the PIM and non-PIM groups. Propensity scores were estimated using logistic regression including all baseline covariates (see S2 and S3 Tables). 1:4 nearest-neighbor matching with a caliper of 0.2×SD[logit(PS)] was applied to maximize power while maintaining balance[30]. The matched dataset was subsequently analyzed using a Cox regression. Results from all Cox regression models were reported as hazard ratios (HRs) and their associated 95% confidence intervals (CIs). The proportional hazards assumption was verified for each model through scaled Schoenfeld residuals, both statistically and visually. Pre-and post-matching, the balance of covariates between groups was assessed by calculating standardized mean differences (SMDs).

Lastly, within the PIM-prescribed population, we compared patients who received only one PIM with those prescribed two or more PIMs to assess whether poly-PIM exposure was associated with higher mortality risk. This subgroup analysis also applied 1:4 PSM (caliper: 0.2) followed by Cox regression.

Proportional hazards assumptions were assessed using scaled Schoenfeld residuals (global and covariate-specific tests). Given the large sample size, visual inspection of smoothed residual plots was prioritized over P-values alone. We utilized SAS (Version 9.4, SAS Institute, Cary, NC, USA) for data management and performed statistical analyses with R software (Version 4.4.1). Statistical significance was set at 5% (p < 0.05).

### Ethical Considerations

This study complied with the Personal Information Protection Act and the healthcare data utilization guidelines by submitting a research proposal to the HIRA to obtain the necessary data. Based on the use of secondary data that preclude personal identification, the study was exempted from Institutional Review Board review (HIRAIRB-2025–051–001); owing to its retrospective design and reliance on anonymized administrative records, informed consent was not required.

## Results

### Baseline Characteristics

Baseline characteristics of the analytic cohort are presented in Table 1. There were no missing values. This study included 943,332 participants, with 15,570 (1.65%) receiving PIM prescriptions. Women comprised a significantly greater proportion of the PIM group (73.44%) compared to the non-PIM group (63.99%) (p < 0.001). Compared with the non-PIM group (72.44 years), the PIM group had a significantly higher mean age (73.37 years) (p < 0.001). For insurance type, the proportion of Medical Aid beneficiaries was higher in the PIM group (10.00%) compared with the non-PIM group (6.57%) (p < 0.001).

**Table 1.** Baseline Characteristics.

| **Characteristic** | **Non-PIM group**<br>**(N = 927,762)** | **PIM group**<br>**(N = 15,570)** | **P-value** |
| --- | --- | --- | --- |
| **Sex (%)** |  |  | <0.001 |
| Female | 593,675 (63.99) | 11,435 (73.44) |  |
| Male | 334,087 (36.01) | 4,135 (26.56) |  |
| **Age (Mean ± SD)** | 72.44 ± 5.98 | 73.37 ± 6.21 | <0.001 |
| **Insurance Type (%)** |  |  | <0.001 |
| National Health Insurance | 866,821 (93.43) | 14,013 (90.00) |  |
| Medical Aid | 60,941 (6.57) | 1,557 (10.00) |  |
| **Type of Medical Institution (%)** |  |  | <0.001 |
| Clinic | 637,024 (68.66) | 11,196 (71.91) |  |
| Long-Term Care Hospital | 3,339 (0.36) | 66 (0.42) |  |
| Hospital | 50,704 (5.47) | 1,041 (6.69) |  |
| General Hospital | 161,307 (17.39) | 2,325 (14.93) |  |
| Tertiary Hospital | 75,388 (8.13) | 942 (6.05) |  |
| **Medical Specialties (%)** |  |  | <0.001 |
| Internal Medicine | 785,373 (84.65) | 11,604 (74.53) |  |
| Neurology | 39,945 (4.31) | 1,754 (11.27) |  |
| Surgery | 18,767 (2.02) | 402 (2.58) |  |
| Orthopedic Surgery | 10,858 (1.17) | 299 (1.92) |  |
| Neurosurgery | 9,588 (1.03) | 332 (2.13) |  |
| Family Medicine | 40,922 (4.41) | 594 (3.82) |  |
| Others | 22,309 (2.40) | 585 (3.76) |  |
| **Emergency Treatment Status (%)** |  |  | <0.001 |
| N | 926,346 (99.85) | 15,384 (98.81) |  |
| Y | 1,416 (0.15) | 186 (1.19) |  |
| **Inpatient/Outpatient Classification (%)** |  |  | <0.001 |
| Outpatient | 923,223 (99.51) | 14,782 (94.94) |  |
| Inpatient | 4,539 (0.49) | 788 (5.06) |  |
| **[Comorbidity]** |  |  |  |
| **Myocardial Infarction (%)** |  |  | 0.009 |
| N | 915,254 (98.65) | 15,398 (98.90) |  |
| Y | 12,508 (1.35) | 172 (1.10) |  |
| **Congestive Heart Failure (%)** |  |  | <0.001 |
| N | 872,121 (94.00) | 14,503 (93.15) |  |
| Y | 55,641 (6.00) | 1,067 (6.85) |  |
| **Peripheral Vascular Disease (%)** |  |  | <0.001 |
| N | 749,849 (80.82) | 11,632 (74.74) |  |
| Y | 177,913 (19.18) | 3,938 (25.29) |  |
| **Cerebrovascular Disease (%)** |  |  | <0.001 |
| N | 798,636 (86.08) | 12,531 (80.48) |  |
| Y | 129,126 (13.92) | 3,039 (19.52) |  |
| **Dementia (%)** |  |  | <0.001 |
| N | 853,335 (91.98) | 13,712 (88.07) |  |

| Characteristic | Non-PIM group<br>(N = 927,762) | PIM group<br>(N = 15,570) | P-value |
| --- | --- | --- | --- |
| Y | 74,427 (8.02) | 1,858 (11.93) |  |
| **Chronic Pulmonary Disease (%)** |  |  | <0.001 |
| N | 721,550 (77.77) | 11,248 (72.24) |  |
| Y | 206,212 (22.23) | 4,322 (27.76) |  |
| **Rheumatologic Disease (%)** |  |  | <0.001 |
| N | 898,234 (96.82) | 14,911 (95.77) |  |
| Y | 29,528 (3.18) | 659 (4.23) |  |
| **Peptic Ulcer Disease (%)** |  |  | <0.001 |
| N | 746,985 (80.51) | 11,325 (72.74) |  |
| Y | 180,777 (19.49) | 4,245 (27.26) |  |
| **Mild Liver Disease (%)** |  |  | <0.001 |
| N | 720,398 (77.65) | 11,772 (75.61) |  |
| Y | 207,364 (22.35) | 3,798 (24.39) |  |
| **Diabetes without complications (%)** |  |  | 0.002 |
| N | 648,320 (69.88) | 11,058 (71.02) |  |
| Y | 279,442 (30.12) | 4,512 (28.98) |  |
| **Diabetes with complications (%)** |  |  | 0.034 |
| N | 830,911 (89.56) | 13,863 (89.04) |  |
| Y | 96,851 (10.44) | 1,707 (10.96) |  |
| **Paraplegia and Hemiplegia (%)** |  |  | <0.001 |
| N | 922,858 (99.47) | 15,423 (99.06) |  |
| Y | 4,904 (0.53) | 147 (0.94) |  |
| **Renal Disease (%)** |  |  | 0.801 |
| N | 905,605 (97.61) | 15,203 (97.64) |  |
| Y | 22,157 (2.39) | 367 (2.36) |  |
| **Any malignancy including leukemia and lymphoma (%)** |  |  | <0.001 |
| N | 873,678 (94.17) | 14,779 (94.92) |  |
| Y | 54,084 (5.83) | 791 (5.08) |  |
| **Moderate or severe Liver Disease (%)** |  |  | 0.031 |
| N | 926,072 (99.82) | 15,530 (99.74) |  |
| Y | 1,690 (0.18) | 40 (0.26) |  |
| **Metastatic solid tumor (%)** |  |  | 0.215 |
| N | 924,925 (99.69) | 15,531 (99.75) |  |
| Y | 2,837 (0.31) | 39 (0.25) |  |
| **AIDS/HIV (%)** |  |  | 0.180 |
| N | 927,626 (99.99) | 15,570 (100.00) |  |
| Y | 136 (0.01) | 0 (0.00) |  |

Analysis of healthcare institution utilization revealed that the PIM group had a higher proportion of visits to primary care clinics (71.91%), whereas the proportion of visits to general hospitals and tertiary hospitals was relatively lower compared to the non-PIM group (p < 0.001). The distribution by medical specialty also showed higher proportions of PIM prescriptions in certain specialties, such as internal medicine, neurology, and surgery (p < 0.001). The proportions of emergency care visits and hospital admissions were also higher in the PIM group (p < 0.001).

Comorbidity analysis indicated that, except for renal disease, most conditions were observed with significantly higher prevalence in PIM users versus non-PIM users. Marked differences were noted in chronic conditions such as peripheral vascular disease, cerebrovascular disease, dementia, chronic pulmonary disease, and peptic ulcer disease (each p < 0.05 or p < 0.001). Moderate to severe liver disease was also more prevalent in the PIM group (p = 0.031). However, metastatic solid tumors and AIDS/HIV did not differ significantly between groups.

Among the participants, 927,762 (98.35%) had no PIM prescriptions, while 1.60% (15,086 individuals) received prescriptions for one type of PIM. Cases involving prescriptions for two or more types of PIMs were very rare, accounting for 0.05% (484 individuals) or fewer (see Table 2). In particular, prescriptions for three or four types of PIMs were extremely uncommon, occurring in only 11 and 1 cases, respectively. Overall, the vast majority of participants did not receive any PIM prescriptions, and multiple PIM prescriptions occurred only in a very small proportion of cases.

**Table 2.**
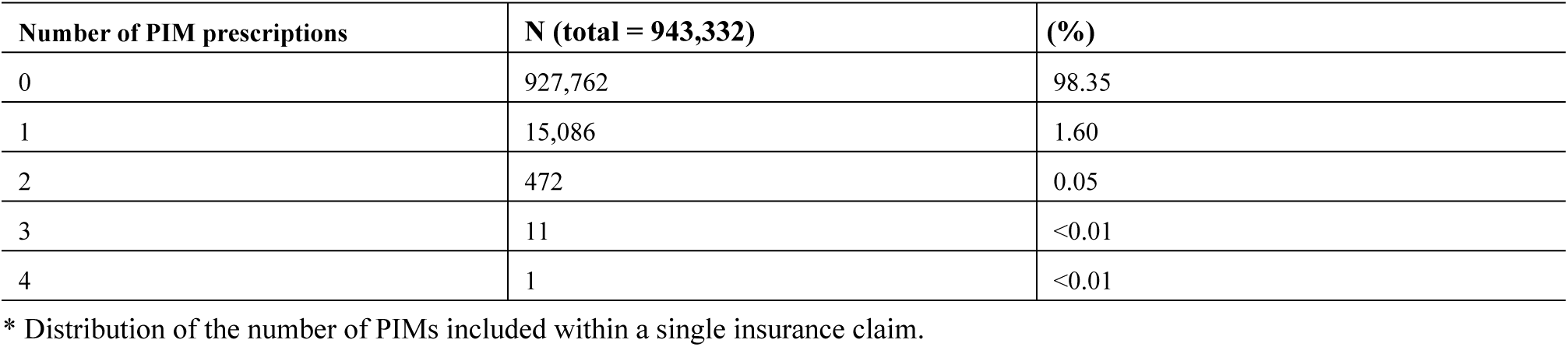
Number of PIM prescriptions.

| Number of PIM prescriptions | N (total = 943,332) | (%) |
| --- | --- | --- |
| 0 | 927,762 | 98.35 |
| 1 | 15,086 | 1.60 |
| 2 | 472 | 0.05 |
| 3 | 11 | <0.01 |
| 4 | 1 | <0.01 |

\* Distribution of the number of PIMs included within a single insurance claim.

Regarding the prescribed medications, the most commonly prescribed PIM was diazepam (60.4%), followed in frequency by amitriptyline hydrochloride (17.0%) and clonazepam (10.1%) (S4 Table presents the proportion of each prescribed medication).

### Basic Analysis and Unadjusted Cox Regression Analysis

During the six-year follow-up (July 1, 2018–June 30, 2024), we observed 2,213 deaths among participants with PIM prescriptions (N = 15,570) and 110,360 deaths among those without PIM prescriptions (N = 927,762). The mean follow-up duration was 5.62 years among those with PIM prescriptions and 5.69 years among participants without PIM prescriptions. The six-year cumulative incidence of death was estimated as 14.2% (95% CI, 13.7–14.8) in the PIM group and 11.9% (95% CI, 11.8–12.0) in the non-PIM group, yielding an absolute risk difference of 2.3 percentage points and a relative risk of 1.19 (log-rank test, χ² = 81.2, p < 0.001). The cumulative incidence function for mortality in the PIM group remained consistently higher throughout follow-up, with progressive divergence over time (see Fig 2).

**Fig 2.**
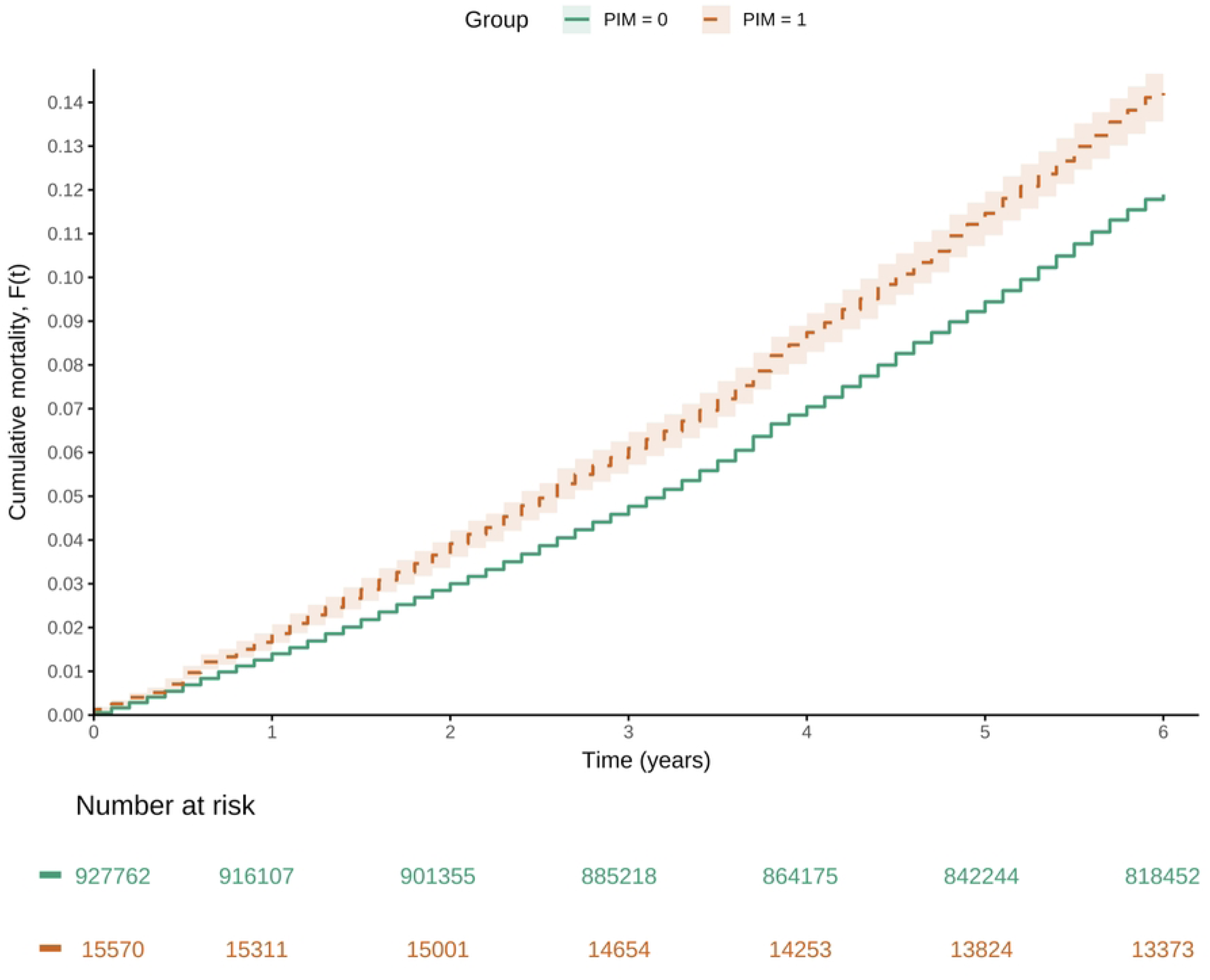
**Cumulative Incidence Function for Mortality in the Entire Cohort**

In the unadjusted univariable Cox regression analysis(detailed results are presented in S5 Table), PIM prescription was associated with a significantly higher risk of death compared with the non-PIM group (HR, 1.21; 95% CI, 1.16–1.27; p < 0.001). By sex, males showed a significantly higher hazard of death than females (HR, 1.75; 95% CI, 1.73–1.77; p < 0.001). Age was also clearly associated with mortality, with the risk increasing for each additional year (HR, 1.14; 95% CI, 1.14–1.14; p < 0.001). Regarding insurance type, Medical Aid beneficiaries showed a significantly elevated mortality risk relative to National Health Insurance subscribers (HR, 1.94; 95% CI, 1.90–1.97; p < 0.001). Patients utilizing long-term care hospitals or hospital-level institutions and above also showed significantly higher mortality risk compared to those visiting clinics (all p < 0.001). Among medical specialties, neurology and neurosurgery were associated with relatively higher mortality risk. Patients receiving emergency care or inpatient treatment also had a markedly increased risk of death (both p < 0.001).

In addition, patients with major comorbidities—including myocardial infarction, congestive heart failure, and dementia—exhibited significantly higher mortality risk (all p < 0.001). Meanwhile, mild liver disease showed no statistically significant association with mortality (p = 0.251) and was therefore excluded from the subsequent adjusted multivariable analysis model.

### Results of Adjusted Cox Regression Model

Using the 11 covariates selected based on prior studies—age, sex, insurance type, myocardial infarction, congestive heart failure, peripheral vascular disease, cerebrovascular disease, rheumatologic disease, diabetes (including with complications), and renal disease—an adjusted Cox regression analysis was conducted (Model 1).

The results demonstrated a significant association between PIM prescription and increased mortality risk (HR, 1.10; 95% CI, 1.05–1.15; p < 0.001; Table 3). The mortality risk for males was about twice that of females (HR, 2.00; 95% CI, 1.97–2.02; p < 0.001). Older age was also significantly associated with higher mortality risk (HR, 1.14; 95% CI, 1.14–1.14; p < 0.001). Regarding insurance type, Medical Aid beneficiaries exhibited an elevated risk relative to National Health Insurance subscribers (HR, 1.44; 95% CI, 1.42–1.47; p < 0.001). Major comorbidities, including myocardial infarction, congestive heart failure, cerebrovascular disease, rheumatologic disease, diabetes, and renal disease, were all related to a significantly higher risk of death (all p < 0.001), whereas peripheral vascular disease did not reach statistical significance (p = 0.176).

**Table 3.** Results of Adjusted Cox Proportional Hazards Regression Model.

| Variable | Adjusted Model 1 |  |  | Adjusted Model 2 |  |  |
| --- | --- | --- | --- | --- | --- | --- |
|  | HR (95% CI) |  | P-value | HR (95% CI) |  | P-value |
| **PIM** (ref. no prescribing) | 1.10 | (1.05-1.15) | <0.001 | 1.02 | (0.98-1.07) | 0.281 |
| **Male** (ref. Female) | 2.00 | (1.97-2.02) | <0.001 | 1.92 | (1.90-1.94) | <0.001 |
| **Age** | 1.14 | (1.14-1.14) | <0.001 | 1.13 | (1.13-1.14) | <0.001 |
| **Medical Aid** (ref. National Health Insurance) | 1.44 | (1.42-1.47) | <0.001 | 1.39 | (1.36-1.41) | <0.001 |
| **Type of Medical Institution** (ref. Clinic) |  |  |  |  |  |  |
| Long-Term Care Hospital | - | - | - | 1.40 | (1.29-1.51) | <0.001 |
| Hospital | - | - | - | 1.25 | (1.22-1.28) | <0.001 |
| General Hospital | - | - | - | 1.27 | (1.25-1.29) | <0.001 |
| Tertiary Hospital | - | - | - | 1.35 | (1.32-1.38) | <0.001 |
| **Medical Specialties** (ref. Internal Medicine) |  |  |  |  |  |  |
| Neurology | - | - | - | 1.13 | (1.10-1.16) | <0.001 |
| Surgery | - | - | - | 1.17 | (1.13-1.22) | <0.001 |
|  | HR (95% CI) |  | P-value | HR (95% CI) |  | P-value |
| Orthopedic Surgery | - | - | - | 1.03 | (0.97-1.10) | 0.306 |
| Neurosurgery | - | - | - | 1.14 | (1.08-1.19) | <0.001 |
| Family Medicine | - | - | - | 0.91 | (0.88-0.94) | <0.001 |
| Others | - | - | - | 1.17 | (1.14-1.22) | <0.001 |
| Inpatient (ref. outpatient) | - | - | - | 1.65 | (1.57-1.74) | <0.001 |
| Myocardial Infarction (ref. No) | 1.34 | (1.30-1.39) | <0.001 | 1.26 | (1.21-1.30) | <0.001 |
| Congestive Heart Failure (ref. No) | 1.47 | (1.45-1.50) | <0.001 | 1.36 | (1.33-1.38) | <0.001 |
| Peripheral Vascular Disease (ref. No) | 1.01 | (1.00-1.02) | 0.176 | 1.01 | (1.00-1.03) | 0.053 |
| Cerebrovascular Disease (ref. No) | 1.42 | (1.40-1.44) | <0.001 | 1.16 | (1.14-1.18) | <0.001 |
| Dementia (ref. No) | - | - | - | 1.68 | (1.65-1.70) | <0.001 |
| Chronic Pulmonary Disease (ref. No) | - | - | - | 1.13 | (1.11-1.14) | <0.001 |
| Rheumatologic Disease (ref. No) | 1.15 | (1.12-1.19) | <0.001 | 1.11 | (1.07-1.14) | <0.001 |
| Peptic Ulcer Disease (ref. No) | - | - | - | 0.96 | (0.95-0.98) | <0.001 |
| Diabetes without complications (ref. No) | 1.30 | (1.28-1.31) | <0.001 | 1.25 | (1.24-1.27) | <0.001 |
| Diabetes with complications (ref. No) | 1.42 | (1.40-1.44) | <0.001 | 1.35 | (1.33-1.37) | <0.001 |
| Paraplegia and Hemiplegia (ref. No) | - | - | - | 1.41 | (1.34-1.48) | <0.001 |
| Renal Disease (ref. No) | 1.86 | (1.81-1.90) | <0.001 | 1.71 | (1.67-1.76) | <0.001 |
| Any malignancy including leukemia and lymphoma (ref. No) | - | - | - | 1.44 | (1.41-1.47) | <0.001 |
| Moderate or severe Liver Disease (ref. No) | - | - | - | 1.64 | (1.49-1.80) | <0.001 |
| Metastatic solid tumor (ref. No) | - | - | - | 3.65 | (3.45-3.86) | <0.001 |
| AIDS/HIV (ref. No) | - | - | - | 1.51 | (1.05-2.19) | 0.028 |

\* Model 1: covariates selected based on previous studies; Model 2: variables selected using the backward elimination method.
† Neither “Emergency Treatment Status” nor the comorbidity variable “Mild Liver Disease” was included in either of the two models.

Model 2 was developed by excluding the mild liver disease variable, which showed no statistically significant association in univariable analysis, and applying the backward elimination method while forcing inclusion of the primary exposure variable, PIM prescription. Backward elimination ultimately retained covariates including sex, age, insurance type, type of medical institution, medical specialty, inpatient status, and major comorbidities such as myocardial infarction, congestive heart failure, cerebrovascular disease, dementia, diabetes, renal disease, moderate-to-severe liver disease, and Metastatic solid tumor (Table 3). In this model, unlike the previous one, PIM prescription did not demonstrate a statistically significant association (HR, 1.02; 95% CI, 0.98–1.07; p = 0.281). Males still exhibited approximately 1.92 times higher mortality risk compared to females (HR, 1.92; 95% CI, 1.90–1.94; p < 0.001). Increases in age and being a Medical Aid beneficiary also remained clearly associated with greater risk of death.

Regarding type of medical institution, mortality risk was substantially higher for patients treated at long-term care hospitals, tertiary hospitals, general hospitals, and hospitals compared to clinics (all p < 0.001). By medical specialty, neurology, surgery, neurosurgery, and other specialties showed higher mortality risk compared to internal medicine (all p < 0.001). Patients with inpatient status had a 65% increase in mortality risk relative to those with outpatient status (HR, 1.65; p < 0.001). Major chronic and severe conditions—including myocardial infarction, congestive heart failure, dementia, diabetes, renal disease, moderate to severe liver disease, and metastatic solid tumors—were all significantly associated with increased mortality risk (all p < 0.05). Interestingly, peptic ulcer disease was inversely associated with a tendency toward lower mortality risk (HR, 0.96; p < 0.001) (Table 3).

In Model 1, PIM use was significantly associated with higher mortality (HR, 1.10; 95% CI, 1.05–1.15). However, in Model 2, which included additional covariates selected via backward elimination, the association was attenuated and no longer significant (HR, 1.02; 95% CI, 0.98–1.07), suggesting potential confounding by unmeasured severity factors.

### Results of Propensity Score Matching

Using the PSM method, the PIM-prescribed group (“Treated”) was matched to the non-PIM group (“Control”) in a 1:4 ratio, resulting in a final matched set of 15,567 individuals in the PIM group and 62,091 individuals in the non-PIM group (caliper = 0.2). In other words, from the original dataset, 865,671 of the 927,762 individuals in the non-PIM group were excluded from matching, and 3 of the 15,570 individuals in the PIM group were excluded, after which the matched dataset was constructed.

Balance diagnostics indicated that matching reduced baseline imbalance across covariates and improved distributional overlap of the propensity scores (see Fig 3).

**Fig 3.**
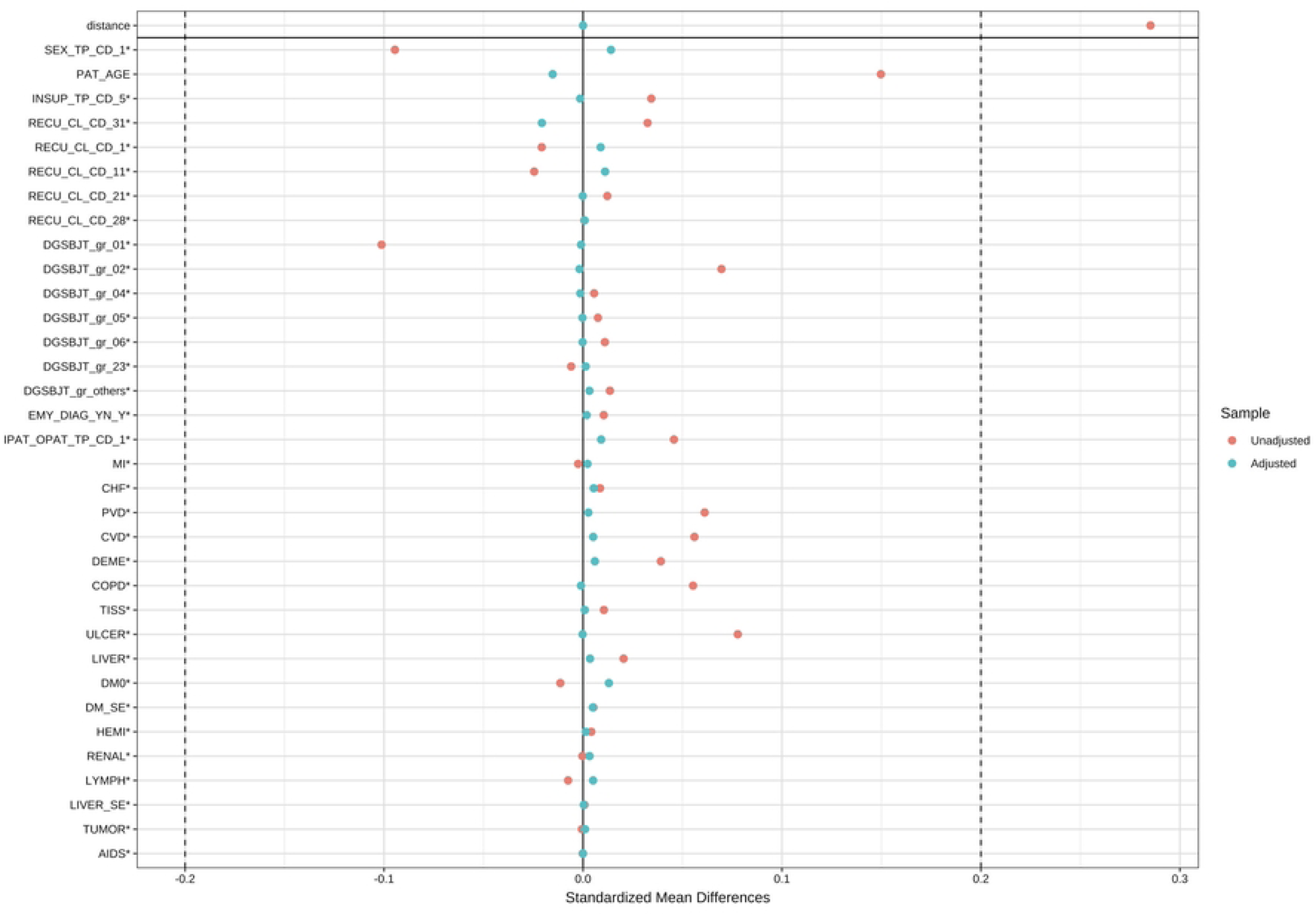
Examining Distributional and Covariate Balance after Propensity Score Matching * Variable names correspond to those described in S2 Table. † Covariate balance assessment based on standardized mean differences before (red) and after (blue) PSM, demonstrating that matching reduced imbalance across all covariates.

Subsequent Cox modeling of the matched sample showed that, similar to Model 1, the PIM-prescribed group exhibited a significantly greater risk of mortality relative to the non-PIM group (HR, 1.12; 95% CI, 1.05–1.19; p < 0.001).

In the 1:4-matched cohort, the cumulative incidence function of mortality increased more steeply in the PIM group than in the non-PIM group, as shown in Fig 4. When evaluated with the log-rank test, the survival distributions of the two groups differed significantly (χ² = 12.3, p < 0.001), suggesting—consistent with the pre-matching analysis—that PIM prescriptions were linked to higher mortality risk.

**Fig 4.**
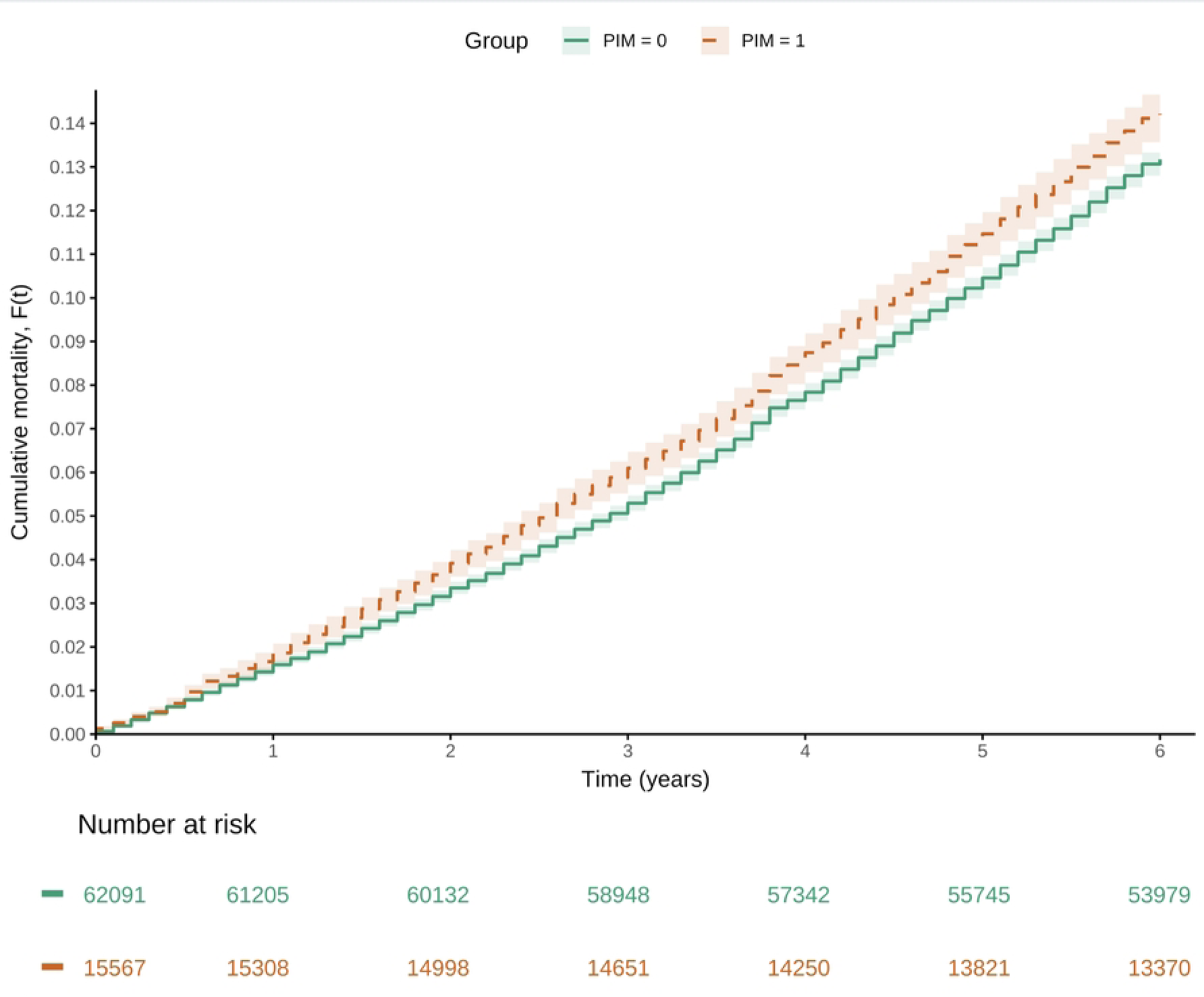
**Cumulative Incidence Function for Mortality in the Matched Cohort**

## Results of Subgroup Analysis

Within the PIM-exposed group, a subgroup analysis compared patients prescribed a single PIM with those prescribed two or more PIMs. Using the PSM method, the group prescribed two or more PIMs (“Treated”; N = 484) and the group prescribed only one PIM (“Control”; N = 15,086) were matched at a 1:4 ratio, resulting in a final matched sample of 483 individuals in the multi-PIM group and 1,892 individuals in the single-PIM group (caliper = 0.2).

Cox modeling of the matched dataset indicated that the group prescribed two or more PIMs had a 1.11-fold higher hazard of events; however, the P-value exceeded 0.05. In other words, the difference in risk between the single-PIM and multi-PIM groups did not reach statistical significance (HR, 1.11; 95% CI, 0.79–1.56; p = 0.544).

### Model Diagnostics

For Adjusted Models 1 and 2, the global Grambsch–Therneau test based on scaled Schoenfeld residuals indicated a statistically significant departure from the PH assumption (p < 0.001). However, given the very large sample size, even small departures can reach statistical significance. In both models, the smoothed residual curves were approximately horizontal around zero with only small, non-systematic fluctuations, suggesting that any time-varying effects were negligible (S6 Table; S1–S2 Fig). By contrast, the PSM model did not reject the PH assumption, and its smoothed residual curves were flat around zero, supporting time-invariant effects (S6 Table; S3 Fig).

## Discussion

In this retrospective cohort study, we evaluated the impact of PIM prescribing on all-cause mortality among older adults (≥65 years) with dyslipidemia. Treating the propensity score–matched analysis as our primary sensitivity analysis, we found a statistically significant elevation in six-year all-cause mortality risk with PIM exposure (HR, 1.12; 95% CI, 1.05–1.19; p < 0.001). The multivariable-adjusted Cox analysis showed a similar association (HR, 1.10; 95% CI, 1.05–1.15; p < 0.001), and the matched-sample log-rank test further supported this pattern. These results suggest that PIM prescribing may have a detrimental impact on the survival of older adults with dyslipidemia.

Our finding that PIM prescribing is associated with increased mortality is consistent with prior meta-analyses[19], and with disease-specific studies (e.g., dementia and kidney disease) reporting similar associations[20, 21]. These findings point to the necessity of careful selection of prescribed medications and routine medication review among older adults, highlighting the importance of more meticulous management strategies that take into account the complex risks associated with comorbidities and polypharmacy.

Notably, model 1 showed a significant relationship between PIM prescriptions and mortality risk, but this did not hold in Model 2. This discrepancy may be explained by the application of the backward elimination method in Model 2, which retained only the most statistically powerful predictors of mortality. The inclusion of variables such as advanced age, severe comorbid conditions, and healthcare utilization patterns-factors known to exert a strong influence on mortality—may have attenuated the independent effect of PIM prescriptions. In the subgroup analysis comparing single-and multiple-PIM prescriptions, no statistically significant difference in mortality risk was detected. However, the multiple-PIM subgroup was relatively small (N = 484), which may have limited statistical power to detect modest differences; thus, the absence of statistical significance should not be taken as evidence of no effect. Alternatively, the incremental effect of receiving ≥2 PIMs over a single PIM may indeed be minimal in this population, or residual confounding from unmeasured variables may have attenuated the association. Taken together, while overall PIM exposure was associated with increased mortality, any additional risk attributable to multiple PIM prescriptions appears less pronounced and warrants confirmation in larger cohorts with more granular clinical and behavioral data.

This study also found that male patients had approximately twice the mortality risk of female patients, that mortality risk increased with age, and that Medical Aid beneficiaries showed greater mortality risk compared to National Health Insurance subscribers. These findings reaffirm that sex, age, and socioeconomic factors are significant determinants of health outcomes in older adults. Therefore, in managing patients with dyslipidemia, a personalized approach that considers each patient’s characteristics and environment—beyond the mere treatment of disease—is essential.

This study has several limitations that warrant recognition. As a retrospective analysis based on claims and mortality data, potential selection bias and information bias cannot be completely ruled out. Prescriptions for non-reimbursed medications were not captured, and important covariates such as marital status, education level, and smoking status could not be included due to data constraints. In addition, although the PIM list in South Korea has continued to expand in recent years—encompassing over 500 products as of August 2025—the number of medications included at the time of cohort assembly in early 2018 was relatively limited, focusing primarily on benzodiazepines and tricyclic antidepressants. This narrower scope may have influenced the observed associations, potentially underestimating the full range of risks associated with PIM use. Future studies incorporating the expanded PIM lists could provide a more comprehensive assessment of prescribing patterns and their clinical implications.

Despite these limitations, the study has notable strengths. By utilizing nationwide claims data linked to official death records, it encompasses a near-complete enumeration of the target population, minimizing selection bias and enhancing the external validity of the results. Furthermore, the large sample size and extended follow-up ensured adequate statistical power to identify meaningful associations between PIM prescriptions and mortality risk in this high-risk group, thereby offering robust evidence that can inform both clinical decision-making and national prescribing policies.

## Conclusion

In this nationwide cohort of older adults with dyslipidemia, baseline exposure to potentially inappropriate medications (PIMs) was associated with elevated six-year all-cause mortality across multiple analytic approaches. These findings support cautious prescribing, routine medication review, and individualized risk–benefit assessments that account for age, sex, comorbidity burden, and socioeconomic context. While the PIM definition used for cohort assembly reflected the national DUR monitoring list in early 2018—which covered a narrower set of drug classes than later iterations—the Korean PIM criteria have since been regularly expanded and refined. Ongoing updates, coupled with real-world outcome studies in dyslipidemia, should guide iterative improvement of prescribing guidance, clinical decision support, and quality indicators. Future research using the expanded PIM lists and richer clinical data (e.g., functional status, over-the-counter medications) will help clarify causal pathways and identify subgroups most likely to benefit from deprescribing strategies. Collectively, these results can support safer medication use and more precise care for older adults with dyslipidemia.

## Supporting Information

S1 File. (DOCX)

S2 File; STROBE and RECORD checklist. (DOCX)

## Author Contributions

Sungju Lee: Study design, Data management, Statistical analysis, Research conduct, Original manuscript preparation

Seungbong Han: Study design, Project oversight, Manuscript review

## Data Availability

The data underlying the findings in this study contain confidential patient information and cannot be publicly released due to the policies of the Health Insurance Review and Assessment Service (HIRA) that restrict external export. However, for researchers who meet the criteria for access to confidential data, the dataset is available from the HIRA Big Data Hub (https://opendata.hira.or.kr/) upon application and approval.

## Acknowledgments

None.

## Data Availability Statement

Data underlying the findings are obtainable from the Health Insurance Review and Assessment Service (HIRA) of South Korea, but access is subject to restrictions. These data were used under license for this study and cannot be publicly released due to policies banning external export. However, researchers meeting the criteria for access to confidential data may obtain the dataset from the HIRA Big Data Hub (https://opendata.hira.or.kr/) after application and approval.

